# Pre-pandemic SARS-CoV-2 potential natural immunity among population of the Democratic Republic of Congo

**DOI:** 10.1101/2021.04.28.21256243

**Authors:** Marc Souris, Léon Tshilolo, Daniel Parzy, Rachel Kamgaing, Destin Mbongi, Baltazar Phoba, Marie-Anasthasie Tshilolo, René Mbungu, Pierre Morand, Jean-Paul Gonzalez

**Author notes:** **Corresponding author** Marc SOURIS. **Funding** “Fond de crise COVID-19”, IRD (Institut de Recherche pour le Développement), Marseille, France.

## Abstract

More than a year after the emergence of COVID-19, significant regional differences in terms of morbidity persist, showing lower incidence rates in sub-Saharan Africa, Southeast Asia, and Oceania. Like SARS-CoV-1 and MERS viruses, SARS-CoV-2 is monophyletically positioned with parental species of chiropteran coronavirus. Furthermore, we observe that the spatial distribution of several targeted bat species (i.e., Coronavirus species hosts) overlaps the distribution of countries with low COVID-19 incidence.

The work presented here aims to test the presence of natural immunity among population with a low COVD-19 prevalence, potentially due to a previous exposure to coronavirus antigens of a virus close related to SARS-CoV-2. To identify such pre-existing immunity, an ELISA serological test was used to detect IgG antibodies targeting main SARS-CoV-2 proteins including: the N-protein, the Spike 1 (S1) protein, the receptor binding domain (RBD) of the S1 protein, the N-terminal domain (NTD) of the S1 protein, and the S2 protein.

A total of 574 sera samples collected before 2019 in the population of the Democratic Republic of Congo (DRC) were tested). 189 control sera from blood donors in France were used as control samples.

The results showed a statistically significant difference between the DRC samples and control samples for all antigens (N, S1, S2, NTD) except for RBD. The percentage of positive samples presenting reactive antibodies for S1 antigen was respectively of 19.2% for RDC versus 2.11% for the control, and of 9.3% versus 1.6% for the S2 antigen.

In conclusion, our data showed that the study population has been potentially exposed to a SARS-CoV-2-like virus antigen before the pandemic in the Central African sub-region. Therefore, it is quite legitimate to think that this prior immunity may be protective and responsible for the observed low prevalence of COVID-19. Moreover, we can assume that this not yet identified SARS-CoV-2-like could be associated to a chiropteran species in close contact with the studied population. In order to confirm the presence of SARS-CoV-2-like virus antibodies and ultimately identify the neutralizing potential for the detected antibodies, our study is underway in other African and Asian countries, where the COVID-19 prevalence is limited.

## INTRODUCTION

### COVID-19 morbidity in the world

More than a year after the emergence of COVID-19, significant regional differences persist, showing the lowest incidence rates in sub-Saharan Africa, Southeast Asia, and Oceania. This trend was observed at the onset of the epidemic and has been confirmed during subsequent waves [WHO 2021].

Many hypotheses have been proposed to explain this situation, including among others: the morbidity and mortality counts are likely to be underestimated in some low- and middle-income countries due to limited epidemiological surveillance and/or public health screening activity; the population of sub-Saharan Africa is younger, with only 2.3% of the population over 65 years of age (whereas people over 65 years of age account for more than three-quarters of the deaths related to COVID-19 in Europe, where this population represents more than 20% of the population); more rural living conditions may reduce the spread of the disease; climatic and environmental conditions unfavorable to the virus and its spread; and/or, the presence of a natural immunity innate (nonspecific) or secondary due to previous contact with a coronavirus close to SARS-CoV-2 and sharing common antigenic profiles. The present work aims to test this last hypothesis.

### Coronavirus and bats

SARS-CoV-2 virus pertains to the Betacoronavirus genus, which includes numerous virus species of Chiropterans. Chiropteran species are also supposed hosts of coronaviruses close to the original strain of SARS-CoV-2 isolated in China [Zhou 2020]. SARS-CoV-1 and MERS viruses are also monophyletically placed with chiropteran coronavirus parental species [Li 2005]. Moreover, in Cambodia and Myanmar, a virus closely related to SARS-CoV-2 was isolated from bat samples (*Rhinolophus Shameli*) collected in 2010 [Mallaparty 2020] [Hul 2021] [Hu 2017]. Globally, it can be observed that the spatial distribution of fruit bats matches the spatial distribution of countries with lower symptomatic circulation of COVID-19, especially in rural populations (Fig. 2). Ecology and behavior of bats, especially fruit bats (e.g.: mass frequentation of fruit orchards, roosting trees close to dwelling) may have favored the direct or indirect contact with humans. Such type of antigenic relationship and acquisition of natural immunity without morbidity, have already been observed with several viruses, including filoviruses and flaviviruses [Gonzalez 2000] [Becquart 2010] [Dos Santos Franco 2019].

**Fig. 1.**
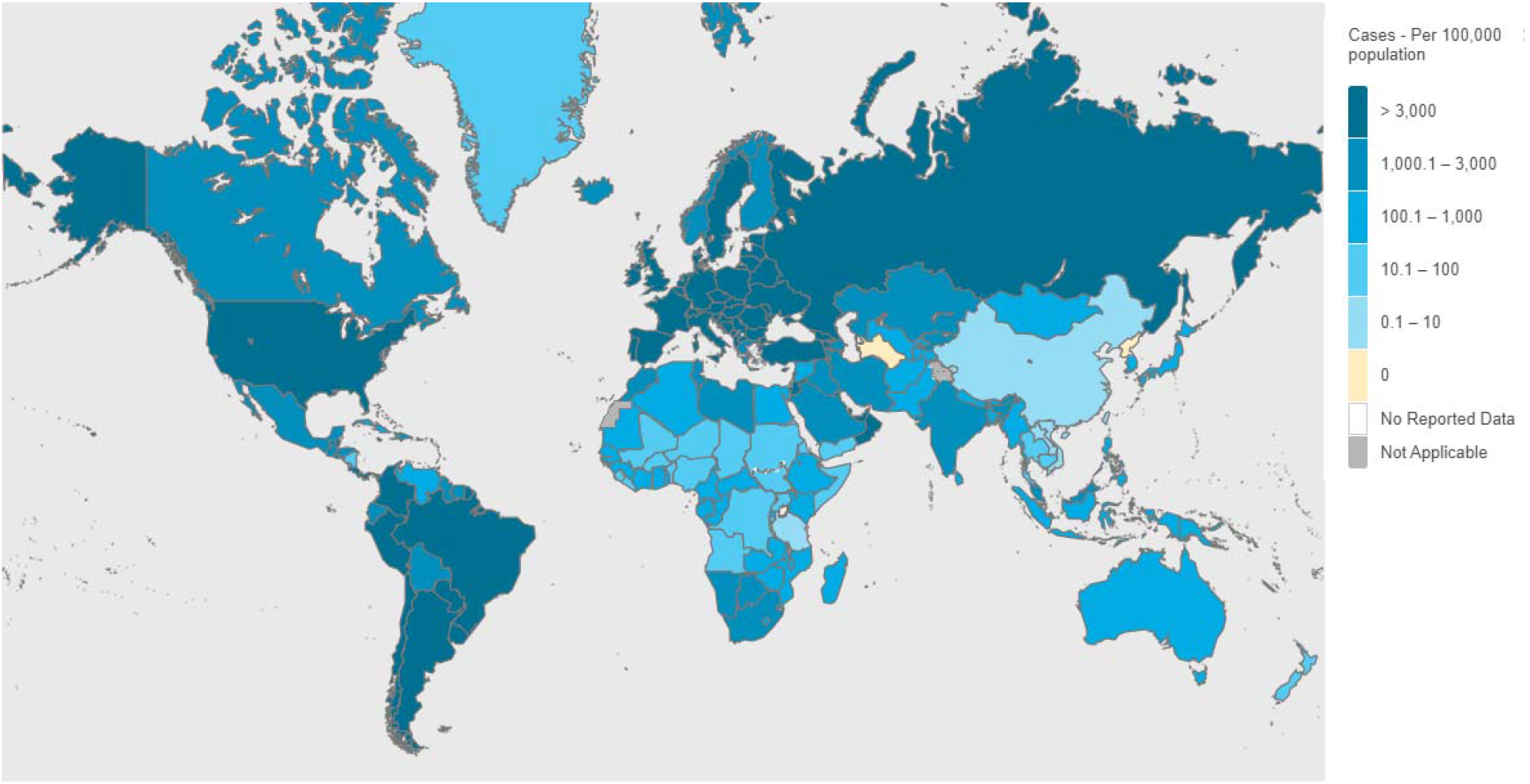
COVID-19 incidence in the World, as for April 19, 2021 (Source: WHO Coronavirus Disease Dashboard, 2021)

**Fig. 2:**
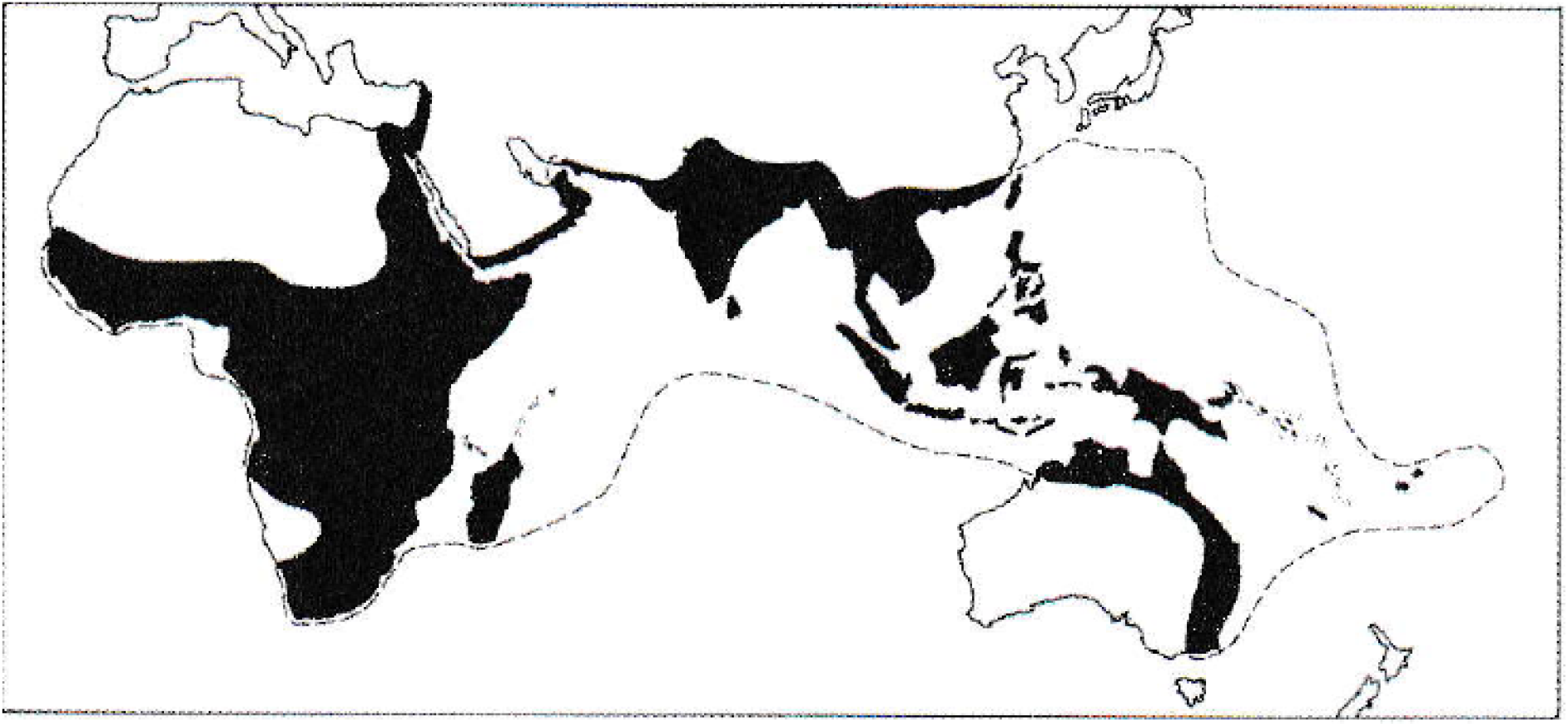
Fruit bats geographic distribution (Source: J. Kingdom, Mammals of Africa, Vol. 4).

### SARS-CoV-2 Antigenic structure

The long genome of Coronaviruses mainly encodes four major structural proteins: spike or S; envelope (E); membrane (M); nucleocapsid (N). The spike-shaped transmembrane glycoprotein (S) on the surface of the virus plays an essential role in virus attachment, fusion, entry, and transmission. It comprises two functional subunits: S1 subunit responsible for binding between the virus and the receptor; and S2 subunit (C-terminal stem) that allows fusion of viral and cellular membranes. The S1 subunit is divided into an N-terminal domain (NTD) and a receptor binding domain (RBD) of the C-terminal region responsible for binding the virus to the host cell (ACE2) receptor binding domain [Lan 2020] [Walls 2020]. The nucleocapsid (N protein) is involved in the packaging of RNA during the externalization of viral particles from the infected cell and is an internal protein of the virus [Wang 2020]. The N protein appears more conserved across Betacoronavirus species than the S protein, while the RBD appears more conserved within the S1 unit. From the point of view of protection (i.e., neutralizing antibodies), there is a strong correlation between the levels of RBD antibodies and the neutralizing antibodies to SARS-CoV-2 in humans [Premkumar 2020]. Also, SARS-CoV-2 genome is closely related to SARS-CoV-1 (79.6% genomic sequence identity), several antibodies covering all structural proteins of SARS-CoV-1 (spike, membrane, nucleocapsid, envelope) have been identified and extensively studied showing cross-reactivity with SARS-CoV-2, as well as partial cross-neutralization of spike antibodies [Bates 2020]. Moreover, sera from SARS-CoV-1 convalescent or S1 CoV-specific animal antibodies can neutralize SARS-CoV-1 infection by reducing S protein-mediated SARS-CoV-1 entry [He 2006]. Finally, SARS-CoV-1 and MERS-CoV show that many fragments (S1-NTD, RBD, S2) of S protein are targets for neutralizing antibody production [He 2006] [Jiang 2020].

The objective of this present study was to identify a pre-existing natural humoral anti-SARS-CoV-2 immunity among population of African countries from sera repository collected several months before the COVID-19 epidemic started. Our work aimed to test the hypothesis of a previous exposure to a SARS-CoV-2 like antigens. We present here our first results, obtained from sera samples collected before January 2020 from people living in the Democratic Republic of Congo (DRC).

## MATERIAL AND METHODS

### Antibody detection

The INNOBIOCHIPS ELISA serological test used, detects the IgG antibodies targeting the N protein, the S1 protein, the RBD domain of the S1 protein, the NTD domain of the S1 protein and the S2 protein. The test values are obtained by optical density reading using a laser reader [INNOBIOCHIPS 2021]. The specificity of the test was evaluated using 25 samples positive for low pathogenic human coronaviruses (229E, OC43, NL63, HKU1) [John 2020]. All 25 samples tested negative for all SARS-CoV-2 proteins used in the ELISA test.

### Control sera collection

The controls sera were obtained by INNOBIOCHIPS company from 189 samples from blood donors collected in Northern France, randomly selected (EFS, Etablissement Français du Sang) and tested negative for SARS-CoV-2 by PCR. These sera collection was used by the manufacturer to define the thresholds of positivity as compared to the sera collected from patients infected by SARS-CoV-2 (PCR test positive). We used this control group to establish thresholds for the absence of SARS-CoV-2-like antibodies with respect to their geographic origin while such blood donors are supposed to have not been in direct or indirect contact with bats. To eliminate the risk of false negatives (samples may came from donors of African or South Asian origin), for each antigen the distribution of control values was modeled. We used negative exponential distribution for the N, S2, RBD, NTD antigens and Weibull distribution for the S1 antigen (fig. 3) and we defined, for each antigen, the PRECOV threshold value corresponding to a probability equal to 0.0002. All control samples with a value for an antigen above the threshold will be considered as false negatives for this antigen. The results for the controls samples are shown in table 1. Among 189 samples, we have 18 samples with at least 1 antigen above the PRECOV threshold: (9.5 %), and 0 sample with at least 2 antigens above the threshold (0 %).

**Fig. 3:**
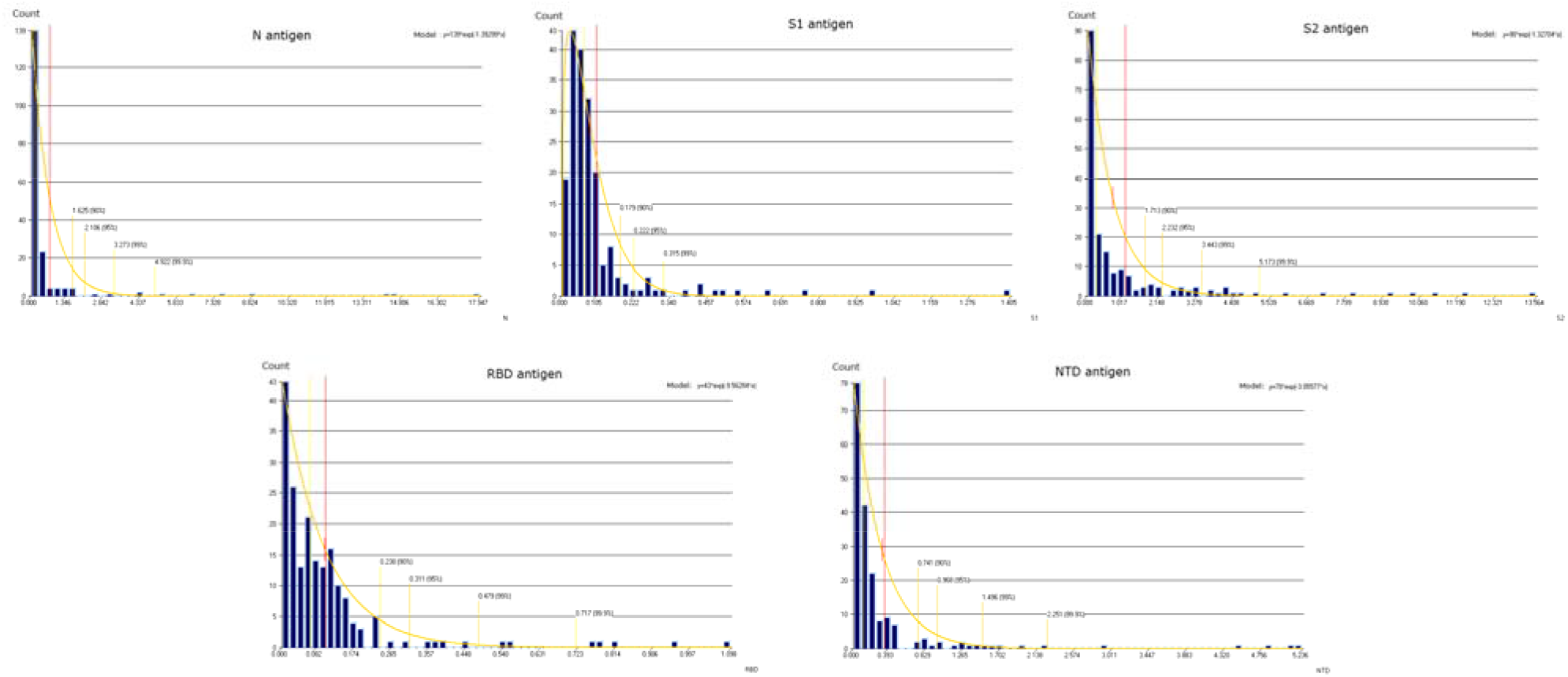
Control sample values distributions for the five tested antigens.

**Table 1.** Values of the INNOBIOCHIPS ELISA test for the controls, PRECOV threshold, and number of samples above the threshold.

| Control (189 sera) | N | S1 | S2 | RBD | NTD |
| --- | --- | --- | --- | --- | --- |
| Min | 0 | 0 | 0 | 0 | 0 |
| Max | 17.95 | 1.40 | 13.56 | 1.10 | 5.24 |
| Mean | 0.695 | 0.106 | 1.125 | 0.105 | 0.353 |
| 1<sup>st</sup> quartile | 0.07 | 0.04 | 0.08 | 0.02 | 0.03 |
| Median | 0.117 | 0.067 | 0.254 | 0.067 | 0.114 |
| 3<sup>rd</sup> quartile | 0.31 | 0.10 | 1.13 | 0.12 | 0.28 |
| Standard Deviation | 2.22 | 0.16 | 2.11 | 0.158 | 0.796 |
| PRECOV threshold | 9 | 0.6 | 10 | 0.6 | 4.5 |
| Number of samples above threshold | 3 (1.6 %) | 4 (2.1 %) | 3 (1.5 %) | 5 (2.6 %) | 3 (1.6 %) |

### Sample sera collection

All 574 samples originated from two sources of sera collected from people living in the Democratic Republic of Congo including: The Monkole Hospital Center biobank (190 samples, collected in 2019); and the ALTADEVA/Monkole biobank (383 samples, collected in 2014 and 2015). The sera from Monkole Hospital Center biobank samples were collected from healthy subjects among the hospital staff, from volunteers, and from young sickle-cell disease patients who are part of a study cohort. The sera, from the ALTADEVA biobank, were collected as part of a study of Plasmodium falciparum chemoresistance, conducted between March 2014 to December 2015 in the city-province of Kinshasa, in the central province of Kongo, and southwestern DRC. All samples were aliquoted and kept frozen as appropriate and each sample had companion data including date of collection, age, sex, and province of origin.

### Data analysis

After calculating the statistical moments and the distribution of the samples’ values for each antigen, several statistical tests and calculations were performed including: For each antigen, comparison of the means (Student’s T-test) and the variances (F-test) between the samples group and the “control” group; Difference between the two groups (samples and controls) considering all the five antigens was tested using Hotelling test; The “control” group was taken as a whole, without excluding the few supposedly false-negative samples; Calculation of the number and percentage of samples considered positive for each antigen (with confidence interval), according to the cutoff value; calculation of the number of positive samples for two or more antigens.

### Ethical Appr ovals

Consent for the study was obtained from all involved. All documents and samples were anonymized. Ethical approval was obtained from the Ethics Committee of the Centre de Formation et d’Appui Sanitaire/Centre Hospitalier Monkole (N/Ref: 01/CEFAMONKOLE/CEL/2013). Blood samples were collected after obtaining informed consent, written in French and in the vernacular language used in Kinshasa and Central Kongo, from each patient or from his or her parent/guardian in the case of minors.

## RESULTS

Antibodies against all five SARS-CoV-2 antigens were detected in the pre-COVID samples, with differential optical density mean value for DRC samples significantly higher than for the control samples, except for RBD antigen The S1 antigen shows the highest percentage of positives: 19.2% for DRC samples versus 2.11% for the control samples. The S2 antigen also shows high rate: 9.3% for DRC samples vs 1.6% for control samples (Table 2). The Hotelling test confirmed a high significant difference (T2 = 35, F = 7, p-value=2.2*10^−6^) when all antigens were considered together.

**Table 2.** Democratic Republic of Congo study samples values (differential optical density). Caption: For each antigen, the table indicate the distribution of values and the number of samples with value above the PRECOV threshold (count, percentage and confidence interval of the percentage). The T-Test p-value indicate the probability of no difference between the mean of DRC samples and the mean of control samples.

| DRC Samples<br>(N= 574) | N | S1 | S2 | RBD | NTD |
| --- | --- | --- | --- | --- | --- |
| minimum | 0 | 0 | 0 | 0 | 0 |
| maximum | 59.09 | 50.28 | 65.58 | 42.17 | 44.68 |
| Mean | 3.01 | 1.05 | 3.43 | 0.29 | 1.11 |
| Standard deviation | 7.72 | 4.32 | 7.90 | 1.85 | 3.76 |
| 1st quartile | 0.16 | 0.088 | 0.196 | 0.000 | 0.047 |
| Median | 0.62 | 0.167 | 0.763 | 0.062 | 0.157 |
| 3rd quartile | 1.92 | 0.396 | 2.852 | 0.182 | 0.578 |
| Number of samples ><br>PRECOV threshold<br>(count, % and C.I.) | 42 (7.3 %<br>[5.2-9.5]) | 110 (19.2 %, [16.0-22.4]) | 45 (9.3 %) [7.1-11.5]) | 33 (5.7 %) [3.8-7.6]) | 32 (5.4 %) [3.6-7.2]) |
| T-Test (Student) | p=3*10<sup>-5</sup> | p=0.001 | p=4*10<sup>-5</sup> | p=0.08 | p=0.003 |

Among the 574 samples, 201 samples reacted at least against 1 antigen above the threshold (35% vs. 9.5% for the controls), while 50 samples reacted at least against 2 antigens above the threshold (8.7% vs 0% for the controls).

## DISCUSSION

Our results are in favor of the hypothesis that some populations in Africa and from other part of the World might be less susceptible to the SARS-CoV2 infection due to a pre-existing immunity triggered by other Betacoronavirus (alias Sarbecoviruses) of animal origin. Also, higher serological cross-reactivity to SARS-CoV-2 in sub-Saharan African regions than in the USA or elsewhere has already been reported and attributed to higher exposure to human coronaviruses (HCoVs) [Mateus 2020]. Although, the hypothesis that infection with other Coronaviruses can induce a consistent level of pre-existing immunity against SARS-CoV2 has been reported for T cells [Le Bert 2020], this is the first time that specific antibodies against SARS-CoV2 proteins in sera collected before the SARS-CoV-2 emergence in China has been reported in Central Africa. Moreover, it has been showed that cross-reactive T cells against SARS-CoV2 can be induced by Common Cold Coronavirus, SARS-CoV1 or eventually other animal betacoronaviruses [Mateus 2020] [Braun 2020] [Le Bert 2020]. Contrary to what was published by Tso et al. [Tso 2020], the specificity of the test used in this work excludes reactions against congenital human coronaviruses (e.g. HCoV-NL63, HCoV-229E) and confirms that the identified antibodies react precisely against the SARS-Cov-2 proteins. The novelty of our finding is that a stronger cross-reactivity in central African regions may come from exposure, not to HCoVs, but to other animal coronaviruses or more specifically ones associated to chiropteran. Also, several betacoronaviruses have been found in horseshoe bats, as well as specific antibody response to these viruses in fruit bats in Africa [Nziza 2020] [Müller 2007].

Based on what we observed that cross-reactivity to S1 and S2, considered specific to SARS-CoV-2, was stronger than for the N protein, considered common to beta-coronaviruses, therefore, it can be assumed that cross-reactivity to S1 and S2 must have been induced by a virus similar – by this spike epitopes – to SARS-CoV-2, rather than by other known human betacoronaviruses. Indeed, The S1 subunit is the least conserved and cross-reactivity cannot be explained by exposure to the known Human CoVs while the N and S2 proteins are the most conserved. Moreover, such cross-reactivity with S1 does not quantitatively reproduced the S1-RBD or S1-NTD data. These apparent discrepancies can be explained by a variation of antibodies affinities to these epitopes due to the structure of the spike or due to a change in their amino acid sequence. Eventually a more consistent response with S1 could be the fact of a non-tested here CTD protein [Van Elslande 2020]. Altogether such apparent discrepancy of antibody response is in favor of an S1 belonging to a Sars-Cov2-like, while all sequence of the antigens included in the commercial ELISA (INNOBIOCHIPS) were entirely based on the reference SARS-CoV-2 strain. It is also important to note that S and N protein sequences are equally divergent among coronaviruses while the S2 subunit is better conserved than the N protein.

We are aware of the limitations of this somewhat pioneering study. The next step is to conduct sensitive investigations of populations potentially exposed to wild animals and to perform back-to-back with the present ELISA test, an essential SARS-Cov-2 neutralization assays to confirm the surprisingly high S1 cross-reactivity and the potential of the antibody to protect against SARS-CoV-2. In addition, most of the samples tested came from rural areas of Central Congo (e.g.: Kimpese of Lower Congo, Kisantu of Western Congo). Furthermore, if it is true that background reactivity in SARS-CoV-2 serological tests is higher, especially in African populations, this may be due not only to widespread circulation/exposure of/to Sarbecoviruses (alias Betacoronavirus) of animals or chiropterans, but to a potential increased cross-reactivity induced by other microorganisms (e.g., malaria, tuberculosis, etc.) as previously observed [Ng 2020] [Stettler 2016] [Welsh 2010].

## CONCLUSION

In conclusion, our study strongly suggests that a SARS-CoV-2-like virus circulated in the Central African sub-region before the pandemic, inducing a cross immunity and eventual cross-protection in the local population. Given the now well-established phylogeny of Betacoronavirus, it is likely that this not yet identified virus could be associated with chiropteran species. Our study is underway in other African and Asian countries, as well as evaluating the neutralizing potential of these pre-pandemic SARS-CoV-2 antibodies.

## Data Availability

N/A

## Acknowledgements

INNOBIOCHIPS Company, for sharing original data and analytical process.

